# IMPACTS OF “AMBIENT NOISE” IN THE EXECUTIVE FUNCTIONS OF PEOPLE WITH SCHIZOPHRENIA

**DOI:** 10.64898/2026.02.13.26346231

**Authors:** Larissa Mirely dos Santos Rodrigues Saraiva, Aline Mendes Lacerda, Amanda Almeida Rodrigues e Silva, Maria Lúcia de Bustamante Simas, Renata Maria Toscano Barreto Lyra Nogueira

## Abstract

Schizophrenia is a severe neuropsychiatric disorder characterized by positive and negative symptoms and cognitive impairments. The present study aimed to investigate the potential interference of ambient noise on the performance of executive function (EF) tasks in individuals with schizophrenia. The sample consisted of 40 participants, divided equally into two groups: a group of individuals with schizophrenia (SchG) and a healthy control group without neuropsychiatric disorders (HC). All participants did three EF assessment instruments: Trail Making Test, Corsi Block Test, and Maze Test. The experimental design included a test-retest procedure with order counterbalancing: half of the sample began the assessment in the noise condition and the other half in the no-noise condition, to control for order and learning effects. The results indicate that ambient noise has a negative impact on the cognitive performance of individuals with schizophrenia. Specifically, the SchG group performed significantly worse on the Maze Test in the noise condition compared to the no-noise condition. These findings contribute to the understanding of the interactions between sensory and cognitive processes underlying the symptoms of schizophrenia. In addition to their theoretical potential, the results have practical implications, as they support the development of intervention strategies and ambiental adaptations that can improve the functionality and quality of life of people with the disorder.

## INTRODUCTION

Schizophrenia is a complex psychiatric disorder characterized by sensory, cognitive, emotional, and social changes that impairs the daily functioning and quality of life of affected individuals. Among the most consistent cognitive deficits are impairments in executive functions (EF), which include skills such as selective attention, working memory, cognitive flexibility, and inhibitory control. These functions are essential for adapting to complex contexts and for making appropriate decisions in everyday and planning situations (Diamond, 2020).

In everyday life, cognitive demands rarely occur in quiet environments. In typical situations — such as in the presence of simultaneous conversations or in contexts with multiple sound stimuli — it is necessary to select and process relevant information while inhibiting background noise. Evidence from studies with healthy individuals indicates that ambient noise can interfere in executive functions and affect performance in tasks that require cognitive control and flexibility (Har-Shai & Zion, 2021).

Individuals with schizophrenia appear to be increasingly vulnerable to exposure to “ambient noise” (Job, 1999; Landon et al., 2016; Stansfeld, 1992; Bustamante Simas et al., 2021). In addition to reporting high sensitivity to noise, these individuals often describe experiences of disturbance and distress associated with this sound exposure (Job, 1999; Landon et al., 2016; Stansfeld, 1992). In this context, Ghazavi et al. (2023) investigated noise sensitivity (NS) by comparing individuals without mental disorders and individuals with schizophrenia, with and without auditory hallucinations. NS was assessed using Schutte’s Noise Sensitivity Questionnaire. The results indicated that people with schizophrenia have greater sensitivity to noise compared to individuals without mental disorders, and this sensitivity is more pronounced in those patients with auditory hallucinations. However, most research on noise sensitivity in schizophrenia is based predominantly on self-report measures, with a shortage of experimental designs or direct objective assessments. Thus, studies that systematically examine the effects of ambient noise on executive performance in this population are still limited, representing a significant gap in the understanding of the functional limitations associated with schizophrenia.

With regard to ambient noise, the so-called cocktail party (CP) stands out, the sound stimulus adopted in this research. The CP can be characterized as structured and dynamic noise, acoustically similar to social environments with multiple simultaneous sound sources such as parallel conversations, sounds of glasses, footsteps, laughter, background music, and other noises resulting from social interactions, composing a complex and variable acoustic field (Bronkhorst, 2015).

As it is a continuous and multi-source noise that is generally irrelevant, CP tends to be perceived as inappropriate and has been associated with negative repercussions on health and well-being, especially in individuals who are more sensitive to noise (Mohammadian et al., 2023). In the literature, this phenomenon is also referred to as the cocktail party problem, a term that emphasizes the attentional demands involved in selecting relevant auditory information amid multiple competing stimuli (Har-Shai & Zion, 2021).

In relation to CP specifically, the literature involving people with schizophrenia has focused predominantly on investigating speech comprehension processes and susceptibility to perceptual intrusions in contexts with multiple competing sound stimuli (Wu et al., 2016; Zheng et al., 2018). The focus of these studies is understand how auditory overload affects the selection and processing of relevant information in acoustically complex environments.

In this context, Wu et al. (2016) conducted a study with individuals with schizophrenia in which some participants were exposed to auditory stimuli (speech) and others to visual stimuli (lip reading). During the presentation of the stimuli, all participants were simultaneously exposed to a CP scenario. The results indicated reduced brain activity in individuals with schizophrenia, especially in the left posterior-inferior temporal gyrus, a region associated with processes such as visual working memory and selective visual attention, suggesting that the presence of multisource noise may impact neural mechanisms underlying cognitive and perceptual processing.

Withing this neurophysiological perspective, Kliuchko, Heinonen-Guzejev, Vuust, Tervaniemi, and Brattico (2016) conducted a combined electroencephalography and magnetoencephalography (M/EEG) study to examine the neural processing of sound characteristics in the central auditory system as a function of individual sensitivity to noise. The findings indicated that high levels of NS are associated with changes in the codification of acoustic attributes and attenuated sound discrimination in the auditory cortex, suggesting less efficient central auditory processing in more sensitive individuals.

There is also converging evidence pointing to structural changes in subcortical regions involved in sensory integration. Morphometric studies described a reduction in the size and volume of the thalamus in schizophrenia. The review by Byne, Hazlett, Buchsbaum, and Kemether (2009) indicates that such volumetric reductions most pronouncedly affect the anterior and mediodorsal thalamic nuclei, as well as the pulvinar—regions critically involved in the modulation and direction of sensory information. More recent investigations(Perez-Rando et al., 2024), corroborate this perspective by suggesting that NS is more related to dysfunctions in central auditory processing than to alterations in the peripheral auditory system.

In addition to anatomical and functional changes in sensory circuits, schizophrenia is recognized as a heterogeneous disorder characterized by significant impairments in different domains of development, particularly cognition (Martínez et al., 2021; Lainscsek et al., 2019). It is important to note that cognitive deficits have a stable and persistent course, continuing independently of the occurrence of psychotic episodes (Howes, Bukala, & Beck, 2024; Tandon, 2024). In other words, although positive and negative symptoms may show partial remission with pharmacological treatment, cognitive deficits are a more lasting component of the disorder, rarely achieving complete remission.

Among the cognitive impairments in schizophrenia (Hons et al., 2021; Xu & Xian, 2023), EF impairment stands out. As mentioned earlier, executive functions are a set of higher cognitive skills that underlie the planning, organization, and execution of complex behaviors in an adaptive, flexible, and goal-oriented manner. They are related to processes such as reasoning, problem-solving, and information processing (Hamdan & Pereira, 2009; Karbach & Unger, 2014). In its contemporary model, Diamond (2020) proposes that EF are organized around three components — working memory, inhibitory control, and cognitive flexibility — whose subfunctions are modulated by executive attention.

Recent and consistent scientific literature on EF in schizophrenia points dysfunction in multiple domains, including working memory, declarative memory, inhibitory control, cognitive flexibility, and attention (Gilmour et al., 2019; Ruiz-Castañeda et al., 2022; Thuaire et al., 2020; Xu & Xian, 2023). These deficits have direct clinical relevance, as thethe functional capacity of individuals with schizophrenia is often compromised. In this regard, Domingos et al. (2015) observed an association between cognitive performance and functional status in people with psychosis. When asked about cognitive limitations in routine, participants reported difficulties in concentration, attention, memory, planning, and following rules. Although all FEs evaluated showed reduced performance, working memory showed the most significant correlation with functional disability.

It should be noted that difficulties in EF are not restricted to the laboratory setting but extend to everyday functioning. Alekseev and Rupchev (2013) demonstrated that executive deficits are directly reflected in daily life, showing that sense of purpose and life planning are related to cognitive flexibility. In their study, patients with greater impairment in cognitive flexibility had greater limitations in daily activities, a more restricted lifestyle, and less motivational engagement in everyday tasks, reinforcing the central role of EF in the autonomy and functionality of people with schizophrenia.

Despite growing interest in the relationship between auditory processing and cognition in schizophrenia, studios with schizophrenic patients evaluating the interference of noise on executive functions is still scarce.

In general, these studies have designs with high internal consistency but limited ecological validity, that is, with little approximation to typical everyday acoustic conditions (Song et al., 2022; Wright et al., 2014).

Considering the possible implications of NS for cognitive performance, Song et al. (2022) examined the effect of different types of noise on working memory in individuals without mental disorders, verifying the negative impact of NS in this domain. Additionally, Wright et al. (2014) in a systematic review analyzed the effects of noise-induced stress — including CP — on the cognitive performance of healthy individuals and discussed that such effects could be potentially more pronounced in people with schizophrenia, due to the combination of higher NS and already established cognitive impairments.

However, few studies directly and objectively assess the interference of ambient noise during the performance of tasks involving complex cognitive processes, such as executive functions, in individuals with schizophrenia.

Considering this gap, the present study proposes exposure to CP noise, an ecological noise, during the execution of executive function tasks frequently used in neuropsychological practice, as it aims to bring experimental research closer to situations experienced in the clinical context.

The choice of topic was motivated, in part, by reports from people with schizophrenia who describe difficulties in performing activities in the presence of “ambient noise”. This is a proposal that has been little explored in the national and international literature, with the potential to broaden the understanding of the interactions between sensory and cognitive mechanisms in schizophrenia, as well as to support intervention strategies and promote quality of life in this population.

Within this context, we hypothesize that individuals with schizophrenia have more impairment in EF performance in the presence of environmental noise compared to individuals without neuropsychiatric disorders. Moreover, we postulate that neuropsychological instruments for clinical use are sensitive to detecting this effect.

## METHOD

### Participants

Forty volunteers aged between 18 and 57 participated in the study, divided into two groups. The Schizophrenia Group (SchG) consisted of 20 individuals diagnosed with schizophrenia, while the Health Control Group (HC) included 20 participants without a diagnosis of neuropsychiatric disorders. Participants were recruited by non-probabilistic convenience sampling. The sample description is shown in Table 1. The study was approved by the Institutional Review Board under protocol 5.571; CAAE: 40058948722.7.0000.5208, ensuring compliance with ethical principles for research involving human subjects.

### Research instruments

The instruments used in this study were: (1) Snellen optotypes for screening visual acuity; (2) semi-structured interviews, one for the HC and another for the SchG, in order to collect social and clinical data from participants; (3) the Trail Making Test (TMT), widely used to assess EF, especially processing speed and cognitive flexibility; (4) the Corsi Block Transfer Test, as an assessment tool for visuospatial working memory; (5) the Maze Test, which assesses reasoning and problem solving; (6) https://youtu.be/SCsZZa1gPqs, similar to that produced at a cocktail party (CP), headphones, an iPod for sound reproduction, pencils, chairs, and a table for the comfort and convenience of volunteers and researchers.

The TMT was designed by Partington and Leiter in 1938 (Llinàs-Reglà et al., 2015) and has a Cronbach‘s alpha of 0.71. The test is administered using pencil and paper, and one of the steps (A) requires knowledge of numbers in ascending order, specifically up to the number 25 (Malloy-Diniz et al., 2016). The first stage, a training/example of the actual task, consists of presenting the numbers 1 to 8. Then, 25 numbers are arranged randomly on a piece of paper and must be identified in ascending order and connected, forming a trail, for example: 1-2-3-4-5-6-7-8… up to 25. The execution time is registered, as well as whether the task was completed.

The Brazilian standardization of the Corsi Block-Tapping Test was carried out by Fonseca et al. (2017), with a Cronbach‘s alpha of 0.71. The test is administered using a wooden board with nine wooden blocks positioned randomly. The test administrator points to a specific sequence of blocks, and the examinee must reproduce it identically. With each new sequence, the number of blocks and the complexity of the task increase. During the test, the number of correct movements reproduced by the volunteers is counted.

The Maze Test is a subtest of the Neuropsychological Assessment Battery (NAB) and is administered using pen and paper, in which seven mazes to be solved are positioned on individual worksheets. The mazes must be completed within a stipulated and timed period. The better the performance on the task, the less time is used and the higher the score. The worse the performance, the longer the time, and the lower the score. Each maze is independent of the previous one. The test administrator explains that each maze has a beginning and an end, and the order should not be changed. The psychometric validation of the test was performed by Pondé et al. (2021) and presented a Cronbach‘s alpha of 0.97.

### Procedures for data collection

Data collection was conducted individually, in a single meeting with each participant, and began with a careful reading and explanation of the Informed Consent Form. After that, the procedure followed the same sequence for all volunteers:

1. Application of the semi-structured interview (average length of 5 minutes);
2. Visual acuity screening test with Snellen optotypes (average length of 5 minutes);
3. Application of the TMT without the interference of CP noise (average length of 5 minutes);
4. Application of Corsi Block-Tapping Test without interference from “cocktail party” noise (average length of 10 minutes);
5. Application of the Maze Test without interference from CP noise noise (average length of 20 minutes).
6. Application of the TMT with interference from CP noise noise (average length of 5 minutes).
7. Application of Corsi Block-Tapping Test with interference from CP noise noise (average length of 10 minutes).
8. Application of the Maze Test with interference from CP noise noise (average length of 20 minutes).

At the end of the procedure, participants were asked if they had difficulty performing the tasks during the noise. The experimental design followed a test-retest method with counterbalanced order, in which half of the sample began the assessment in the noise condition and the other half in the no-noise condition. This approach aimed to control potential ordering and learning effects, increasing the methodological robustness of the study.

### Data analysis

The TMT data were obtained from the task completion time, as the longer the time, the worse the test performance. In the Corsi Block-Tapping Test, the raw scores corresponding to the number of correct answers were analyzed, as the higher the score, the better the test performance. Finally, in the Maze test, which uses the test completion time as a measure, a score was assigned according to that time — the shorter the time, the higher the score; therefore, the higher the score, the better the performance.

For these neuropsychological tests, descriptive analyses were performed — means and standard deviations were calculated for each group, and graphs were constructed. A 2 × 2 MANOVA with three dependent variables was conducted. Assumptions of multivariate normality, homogeneity of covariance matrices, and absence of multicollinearity were met. Wilks’ Lambda indicated a significant multivariate effect, Λ = .39, *F*(6, 33) = 3.57, *p* = .008.

The sociodemographic data were analyzed using descriptive statistics, and only age was tested using a nonparametric test to compare groups (Mann-Whitney U test), since it is a discrete variable.

## RESULTS

The sample of both groups consisted of 13 male participants and 7 female participants. The mean age of the Schizophrenia Group (SchG) was 33.45 years (SD = 8.35), while that of the Control Group (HC) was 33.90 years (SD = 9.00), indicating a similar age distribution between the groups.

Regarding education, it was noted that 60% (n = 12) of HC participants had completed high school, whereas only 35% (n = 7) of SchG participants had achieved a similar level of education. In addition, 30% (n = 6) of SchG participants had not completed elementary school, in comparison to 15% (n = 3) of HC participants who had not met this educational level.

Concerning noise perception, 70% (n = 14) of SchG volunteers reported discomfort with CP noise, whilst no HC participants complained about this matter, highlighting perceptual differences between the groups.

Among the medications used by SchG, we can highlight that the most commonly used was haldol, 60% (n=12), followed by promethazine, used by 40% (n=8) of participants; biperiden by 35% (n=7), clonazepam and risperidone were used by 30% of participants. Only one volunteer was unable to report their current medication, and the information was also not included in the medical records.

Regarding the comparisons between SchG and HC, Figure 1 shows the results of the TMT, displaying the average task execution time in each group in the conditions with and without noise. In both conditions, SchG had longer execution times than HC, indicating greater difficulty in performing the task. The differences between the groups were statistically significant in both conditions, as evidenced by the *p* values (*p* < 0.05 in the no-noise condition and p = 0.01 in the noise condition), demonstrating a consistent effect of the diagnosis on performance, regardless of the presence of ambient noise

**Figure 1.**
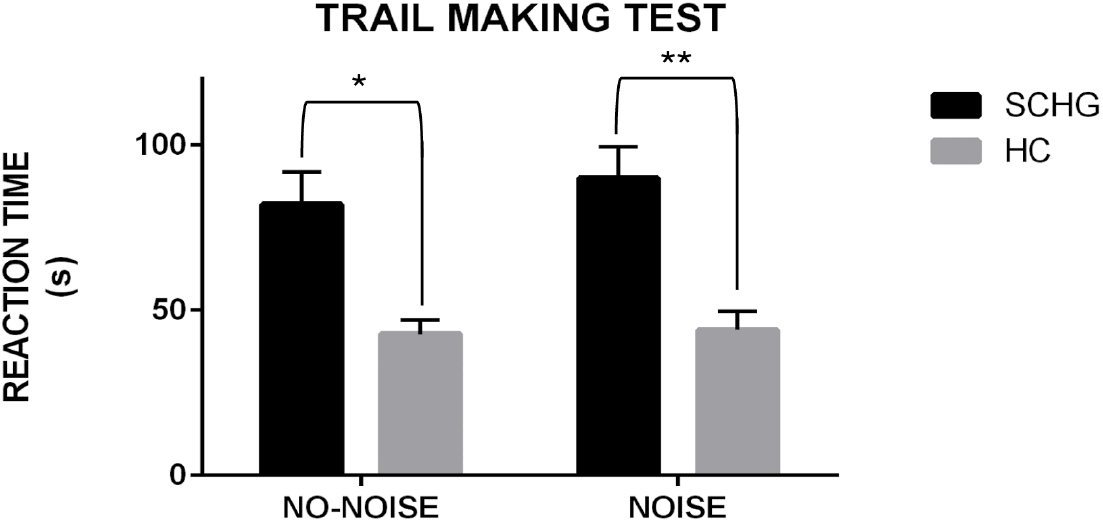
Average reaction time to complete the TMT per group. Note. * p < 0.05 ** p < .001; *ANOVA/ MANOVA*. Caption: The figure shows a graph whose y-axis represents the no-noise and noise conditions. For each condition, there are two vertical bars: a black one, representing the reaction time of the SchG, and a gray one, representing the reaction time of the HC. The x-axis represents the reaction time. In both conditions, the black bar is longer than the gray bar. In each condition, there is a bracket indicating the difference between groups with one and threeasterisks each. This graph describes the results for the Trail Making Test (TMT).

Referring to the Corsi Block-Tapping Test, as shown in Figure 2, the SchG group performed worse than the HC group, as evidenced by a lower average number of correct responses in both conditions. This difference between groups was statistically significant, with p < 0.05 only in the noise condition.

**Figure 2.**
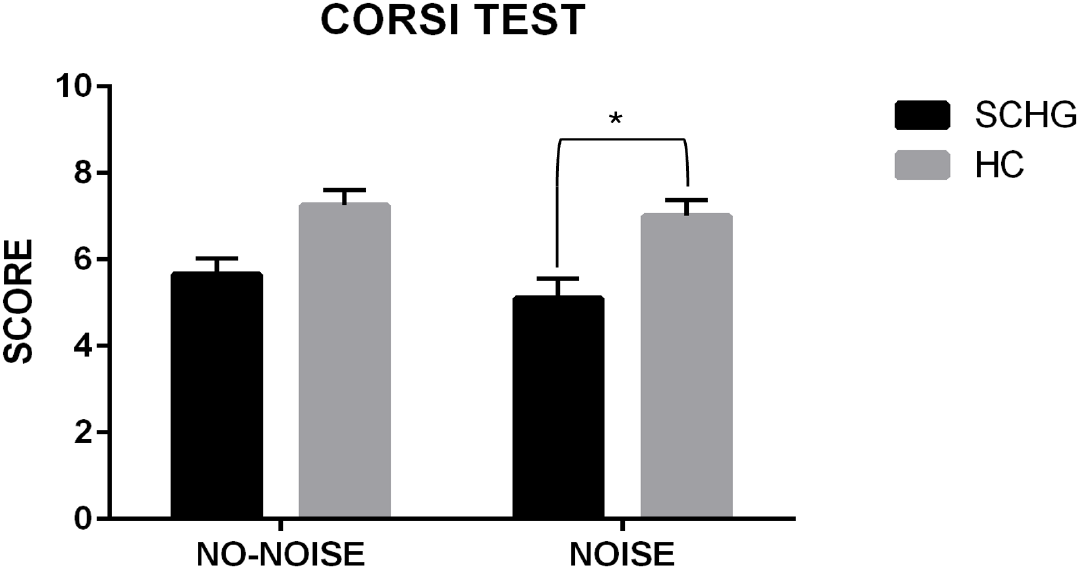
Average number of correct responses in the Corsi Block-Tapping Test by group. Note. * p < .05; *ANOVA/ MANOVA*. Caption: The figure shows a graph whose y-axis represents the no-noise and noise conditions. For each condition, there are two vertical bars: a black one with the SchG score and a gray one with the HC score. The x-axis represents the score. In both conditions, the gray bar is larger than the black bar, indicating that the HC obtained a higher score than the SchG. There is a bracket indicating the difference between groups with one asterisks each for noise conditions. This graph describes the results for the Corsi Block-Tapping Test.

Figure 3 illustrates a graph describing the average scores on the Maze Test for the groups, also according to noise conditions. The figure shows that the SchG group again scored lower than the HC group, i.e., patients performed worse on the task. This difference between the groups was not significant, but the SchG scores differed in noise and no-noise conditions, with p < .05.

**Figure 3.**
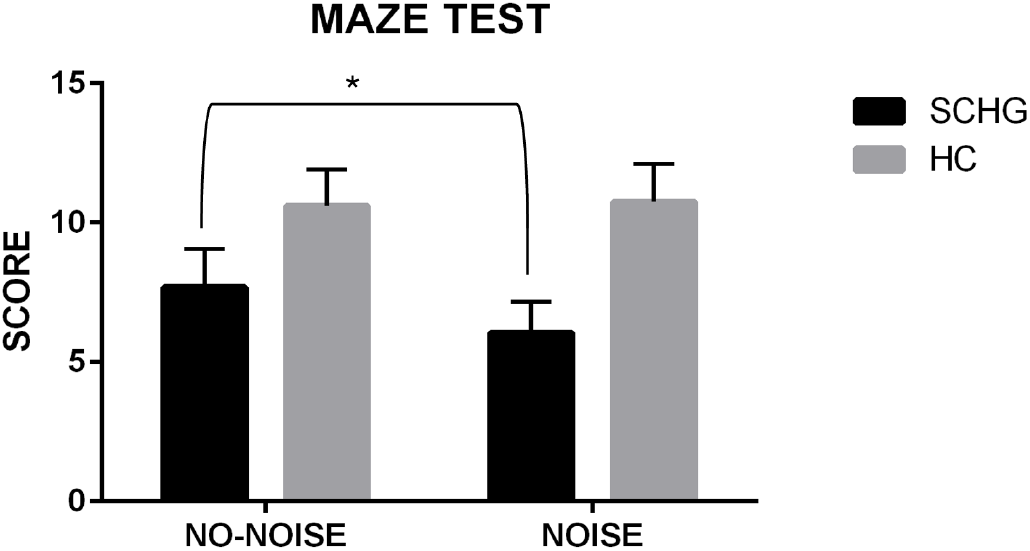
Maze test score by group. Note. * p < .05; *ANOVA/ MANOVA*. Caption: The figure shows a graph whose y-axis represents the no-noise and noise conditions. For each condition, there are two vertical bars: a black one with the SchG score and a gray one with the HC score. The x-axis shows the score. In both conditions, the gray bar is larger than the black bar, indicating that the HC obtained a higher score than the SchG. There is a bracket connecting the two conditions of the SchG (noise and no-noise) with one asterisk, showing a significant difference between the two conditions. This graph describes the results for the Maze Test.

In general, it was observed that the average performance in noise condition was lower in both groups and in all tests. However, a statistically significant difference was only found in the Maze Test for SchG, while HC showed no significant variation in performance between noise and no-noise conditions in any of the three tests evaluated. These results indicate that the presence of ambient noise has a more pronounced impact on individuals with schizophrenia.

## DISCUSSION

The primary aim of this study was to examine the potential interference of noise/cocktail party (CP) conditions on executive function (EF) task performance in individuals with schizophrenia (SchG).

The findings further support existing literature by demonstrating that individuals with schizophrenia exhibit poorer performance on executive function tasks compared to individuals without the disorder. Moreover, the healthy control (HC) group did not show significant performance decrements under noise interference. That is, CP conditions did not adversely affect HC performance on the Trail Making Test, Corsi Test, or Mazes Test in HC.

As for SchG, we observed worse average performance in the three EF tests in the presence of noise/CP. However, there was only a significant difference in the Maze Test, i.e., SchG performed worse in the Maze task with noise interference, a result that responds to the primary objective of this research, and corroborate our hypothesis that noise/CP interferes with the performance of EF in people with schizophrenia. As qualitative data, the SchG‘s self-report of feeling bothered by noise/CP corroborates the studies by Bustamante Simas et al. (2021), Ghazavi et al. (2023), Job (1999), Landon et al. (2016), Mohammadian et al. (2023), and Stansfeld (1992). From a neurophysiological point of view, our study can also be associated with changes in thalamic regions described by Perez-Rando et al. (2024). It is possible that the thalamus is not filtering noise adequately, leading to altered cognitive processing.

Regarding the comparison of noise and no-noise conditions, we again highlight that HC showed no difference between conditions, however SchG showed worse performance in all tests when performed in the presence of noise, and only the Maze Test showed a significant difference.

As mentioned earlier, the results of this study suggest that CP interferes in performance on executive function tasks in people with schizophrenia, as do studies such as those by Song et al. (2022) and Wright et al. (2014) found that the presence of noise, such as the CP and NS, affects cognitive performance in different ways, especially in working memory, attention, and functional capacity. These considerations corroborate the results of the present study.

We can speculate that this significant difference only in the Maze Test was due to the longer time required to complete it. While the TMT took an average of approximately 2 minutes to complete, the Maze Test took an average of 40 minutes. Perhaps the presence of noise/CP interferes with the execution of longer tasks that require more sustained attention.

Another explanation for this result concerns the presence of extrapyramidal symptoms, such as acute dystonia and pseudoparkinsonism (Owen et al., 2012) in patients with schizophrenia, since most of them use typical antipsychotics that produce cholinergic action related to the presence of these symptoms, and all the tests used recruited motor responses. The Corsi Block-Tapping Test required volunteers to move their hands to touch the sequence of cubes presented, the TMT required them to trace a path with a pencil, and the Maze Test, in addition to requiring them to trace a path with a pencil, had a very narrow space for tracing. Thus, we can assume that the need for more sophisticated fine motor skills in the Maze may have been influenced by extrapyramidal symptoms.

Throughout our results, we describe the comparison of performance in EF tasks between people without mental disorders and people with schizophrenia, respectively in the Corsi, Trails, and Maze Tests. We observed worse performance by the SchG in these tasks in both conditions (noise and no-noise), and the Trail Making Test showed the greatest difference between the groups. These data corroborate previous studies, as this test has been shown to be very sensitive in assessing cognitive impairments in people with schizophrenia (Gilmour et al., 2019; Ronenwett, 2016; Ruiz-Castañeda, et al., 2022; Thuaire et al., 2020; Xu & Xian, 2023).

Finally, we would like to highlight some limitations of our study, among them: 1) we did not reach the sample size of 68 participants for medium power; 2) the voices in the CP were in English, and we might have had more difficulties with both groups if the voices were in the participants’ language (Brazilian Portuguese).

## CONCLUSIONS

This study investigated possible effects of noise/CP on performance in EF tasks in people with schizophrenia. Three neuropsychological tests were administered to two distinct groups, one consisting of people with schizophrenia (SchG) and one consisting of people without a diagnosis of mental disorders (HC). The results indicate that noise/CP interferes in the cognitive performance of people with schizophrenia, as the SchG showed significantly worse performance on the Maze Test in the noise/CP condition when compared to performance in the no noise/CP condition.

The results suggest that people with schizophrenia not only feel more bothered by CP, but that it also seems to impair their performance on longer tasks that require EF with finer motor coordination, suggesting greater vulnerability of their executive functioning to sensory interference.

It is suggested that further studies be conducted in this direction, using a greater number of tests that assess EF, as well as more extensive tests that require more time to administer.

## Data Availability

All data produced in the present study are available upon reasonable request to the authors

https://www.dropbox.com/scl/fi/qw2aydrqmp1iedplckts1/Pr-print.pdf?rlkey=gtvpjfs2ioi3sg0003hex34th&st=u5ut5a51&dl=0

## Acknowledgements

We express our gratitude to the professionals and service users of the CAPS in the Agreste region of Pernambuco, as well as to the professionals of the Brazilian Unified Health System (Sistema Único de Saúde – SUS - Brasil)

## Notes

### Competing Interest Statement

The authors have declared no competing interest.

### Funding Statement

This study did not receive any funding

### Author Declarations

Universidade Federal de Pernambuco, Comite de Etica em Pesquisa do Centro de Ciencias da Saude da Universidade Federal de Pernambuco. The study was approved by the Institutional Review Board under protocol 5.571; CAAE: 40058948722.7.0000.5208, ensuring compliance with ethical principles for research involving human subjects.

